# Utility of Cloth Masks in Preventing Respiratory Infections: A Systematic Review

**DOI:** 10.1101/2020.05.07.20093864

**Authors:** Agnibho Mondal, Arnavjyoti Das, Rama Prosad Goswami

## Abstract

**Background:** Using face masks is one of the possible prevention methods against respiratory pathogens. A number of studies and reviews have been performed regarding the use of medical grade masks like surgical masks, N95 respirators etc. However, the use of cloth masks has received little attention.

**Objectives:** The purpose of this review is to analyze the available data regarding the use of cloth masks for the prevention of respiratory infections. We intended to use data from both clinical and non-clinical studies to arrive at our conclusion.

**Methods:** We used PubMed, Cochrane Library and Google Scholar as our source databases. Both clinical and non-clinical studies, which had data regarding the efficacy of cloth masks, were selected. Articles not containing analyzable data including opinion articles, review articles etc. were excluded. After screening the search results, ten studies could be included in our review.

Data relevant to our objective was extracted from each study including clinical efficacy, compliance, filtration efficacy etc. Data from some studies were simplified for the purpose of comparison. Extracted data was summarized and categorized for detailed analysis. Qualitative synthesis of the data was performed. But the heterogeneity between the studies did not allow for a meta-analysis.

**Discussion:** The review was limited by a lack of sufficient clinical studies. Lack of standardization between studies was another limitation.

Although cloth masks generally perform poorer than the medical grade masks, they may be better than no masks at all. Filtration efficacy varied greatly depending on the material used, with some materials showing a filtration efficacy above 90%. However, leakage could reduce efficacy of masks by about 50%. Standardization of cloth masks and appropriate use is essential for cloth masks to be effective. However, result of a randomized controlled trial suggest that they may be ineffective in the healthcare setting.

## Introduction

One of the commonly employed methods to prevent the spread of any pandemic caused by air-borne respiratory pathogens is use of physical barrier methods like protective clothing and mask usage. The primary method of transmission of respiratory pathogens especially viral infections is via droplets. Physical barriers like masks may have some benefit in preventing such infections by preventing the droplet spread from person to person.

At the moment of conducting this review, the COVID-19 pandemic has been sweeping through the globe and a number of preventive measures have been implemented including but not limited to social distancing, hand hygiene, respiratory etiquette etc. Some countries have even recommended the use of cloth masks for the general population.^1^ However, a closer look at the available evidences is necessary regarding the matter of cloth masks.

A 2011 systematic review and meta-analysis in the Cochrane Database for Systematic Reviews^2^ analyzed results of 67 studies on the effect of physical interventions to prevent the spread of respiratory pathogens. Data from nine case control studies suggested that physical barriers were effective in this regard. Both surgical masks and N95 masks were found to be effective, however there was no information about the efficacy of cloth masks.

In the time of a pandemic there is a noted scarcity of resources, which includes *medical grade masks* like surgical masks and other respirators like N95. In such times of crisis, the policymakers especially in the community setting may seek supplementation of these equipments. Hence, whether cloth masks may be used in the place of medical grade masks, needs to be answered.

Cloth masks are different from medical grade masks. They are not standardized and there is no standard evidence based guidelines for their use in the context of preventing disease transmission. There is a severe dearth of evidence regarding the efficacy of cloth masks in preventing the transmission of respiratory pathogens. Moreover, due to their much dissimilarity with the medical grade masks, we cannot extrapolate the evidence from one to the other. Hence, although the systematic review of Jefferson T et al.^2^ concluded that use of masks is likely to be effective in preventing disease transmission, the same cannot be said about the cloth masks without further evidence.

Therefore, to address this research gap, our study aims to analyze the available evidences to find the utility of cloth masks in preventing respiratory pathogen transmission. Since only a handful of clinical studies are available we will also take into account the non-clinical studies which have a direct clinical implication in this matter.

## Methods

### Study Question

The question we seek to answer is stated as follows

- Is cloth mask useful in preventing respiratory infection?

### Data Sources and Searches

We reported our study in compliance with Preferred Reporting Items for Systematic Reviews and Meta-Analyses (PRISMA) guidelines.^3^

We searched the following databases including PubMed, the Cochrane Library and Google Scholar. We used the following search terms

- PubMed and Cochrane Library- *(mask OR masks) AND (cloth OR fabric OR homemade OR home-made OR (home AND made) OR improvised)*
- Google Scholar- *allintitle: mask AND (cloth OR fabric OR homemade OR home-made OR improvised)*

Two authors, AM and AD, performed the search independently and duplicate results were removed.

The results were then evaluated for eligibility by two authors independently, first by the titles, then by the abstracts and lastly by the full texts. References from each included study were also searched for further relevant studies. The result from each author was compared and any disparity was resolved through discussion among all three authors.

### Study Selection

The studies meeting the following criteria were considered for inclusion

- Studies evaluating efficacy of cloth masks in the clinical settings
- Studies evaluating compliance of the subjects to wearing cloth masks
- Studies evaluating filtration efficacy of cloth masks
- Studies evaluating microscopic characters of the cloth masks

We excluded the following types of papers

- Papers on opinions, hypothesis, case reports, case series, letters and editorials
- Review articles
- Non-experimental studies such as mathematical modelling
- Papers with unavailable full text

We did not set any time, geographic or language restriction for search results. The translated entries in the databases and online translator tools were used for non-English studies. All studies published till the date of search were considered for inclusion.

### Data Extraction and Quality Assessment

Data from individual studies were extracted for evaluation, including the clinical outcomes, risk ratio of developing respiratory illness, rate of compliance, filtration efficacy and microscopic characteristics. In case of the papers, having only graphical representation of the data, a graph digitizer was used to extract the necessary data. Data from some studies were simplified to make them comparable to other studies.

### Assessment of Risk of Bias

The included randomized controlled trials were assessed for risk of bias using the Risk of Bias Tool version 2 by the Cochrane Collaboration.^4^ Two authors, AM and AD, performed the assessments independently and any disagreement was resolved by discussion.

### Data Synthesis and Analysis

The extracted data was summarized and categorized based on different aspects of the research question. Tabulation of data was done when possible. The data from the studies were then compared to each other and analyzed to form a conclusion.

## Results

### Included Studies

The literature search was performed on May 2, 2020 according to the PRISMA protocol (Figure 1). The PubMed search yielded 143 results in total, whereas Cochrane Library and Google Scholar produced 140 and 24 results respectively. A total number of 293 studies remained after removing duplicates.

**Figure 1:**
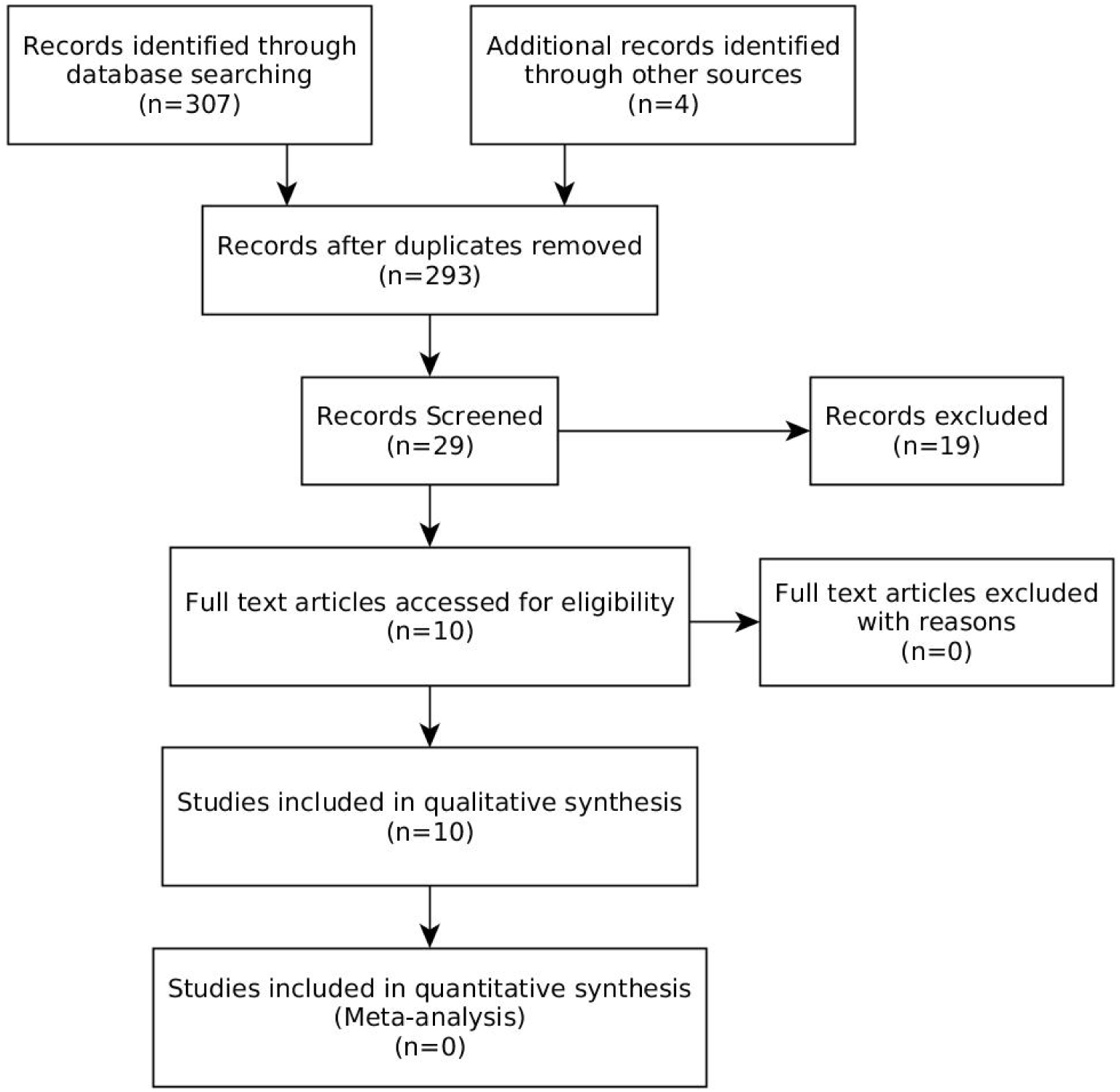
PRISMA statement

After screening, ten studies met our inclusion criteria. Among them two papers were randomized controlled trials and eight were non-clinical studies. The randomized controlled trial comparing efficacy of cloth masks with that of surgical masks in preventing infections was performed in Vietnam.^5^ The second paper used the data from the first trial to assess the compliance of the healthcare workers to cloth masks and surgical masks.^6^ Seven of the non-clinical studies measured filtration efficacy of cloth masks^7-13^ and the remaining study analyzed the microscopic properties of cloth masks.^14^

Risk of bias assessment of the randomized controlled trials is provided in table 1. However, a meta-analysis could not be performed due to a lack of homogeneity of the available data.

**Table 1:**
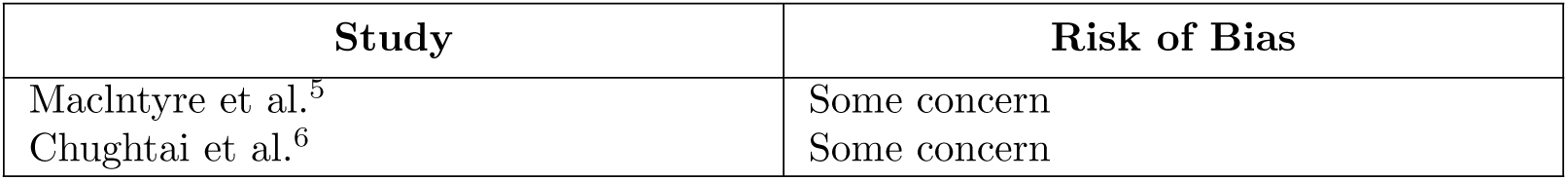
Risk of bias assessment

| Study | Risk of Bias |
| --- | --- |
| MacIntyre et al.<sup>5</sup> | Some concern |
| Chughtai et al.<sup>6</sup> | Some concern |

### Clinical Efficacy

Only one randomized controlled trial evaluated cloth masks for their efficacy in preventing infection by respiratory pathogens in comparison to surgical masks. This randomized controlled trial^5^ by Maclntyre et al. had three arms, surgical masks, cloth masks and standard practice. The total number of participants were 1607, all of them healthcare workers. The study found that after four weeks of follow up, the cloth masks performed poorer than the surgical masks in preventing influenza like illness. The relative risk of influenza like illness was 13.25 (95% confidence interval 1.74 to 100.97) compared to the surgical masks on intention-to-treat analysis. However, there was no appreciable difference between cloth masks and surgical masks regarding laboratory confirmed viruses (relative risk 1.66, 95% confidence interval 0.95 to 2.91).

### Compliance

The compliance to cloth masks in comparison to surgical masks was assessed by Chughtai et al.^6^ using the data from the previous randomized controlled trial. Compliance was defined as the use of designated masks for 70% of the time while on hospital duties. The authors reported a compliance rate of 56.8% in the cloth mask arm and 56.6% in the surgical mask arm. The multivariate analysis showed a relative risk of 1.02 (95% confidence interval 0.97 to 1.08) which indicates that compliance in both groups was almost identical.

However, they also failed to show any association between compliance and the efficacy in preventing infections.

### Filtration Efficacy in Laboratory Setup

Seven of the included studies had evaluated the filtration efficacy of different types of cloth masks (Table 2). Four of these studies were conducted in laboratories with mechanical particles or droplet generators. These particles were passed through the mask under examination and filtration efficacy was calculated depending on the percentage of particles blocked by the mask. Two studies used healthy volunteers. One of them^8^ placed the detector under the mask while it was worn by the subject and counted the ambient particles passing through. In the other study^10^ the volunteers coughed into a box with or without masks and the microbes inside the box were then cultured to estimate the filtration efficacy.

**Table 2:** Filtration efficacy reported by the included studies

| Study | Material | Particle | Filtration efficacy(%) |  |  |
| --- | --- | --- | --- | --- | --- |
|  |  |  | Cloth mask | Surgical mask | N95 mask |
| Sande et al.<sup>8</sup> | Tea cloth | Ambient air particles | 29 - 74 | 55 - 88 | 74 - 99.5 |
| Rengasamy et al.<sup>9</sup> | 5 types | 20 - 1000 nm (polydisperse) | 10 - 26 |  | 99.9 |
| Davies et al.<sup>10</sup> | 10 homemade masks | Expelled droplets during cough | 79 | 85 |  |
| MacIntyre et al.<sup>5</sup> | Cloth mask | Sodium chloride particles | 3 | 56 | 99.9 |
| Shakya et al.<sup>11</sup> | 3 types | PSL >1 $\mu\text{m}$ | 48 - 80 | >97 | 67 - 95 |
| | | PSL <1 $\mu\text{m}$ | 15 - 86 | 63 - 69 | 77 - 88 |
|  |  | Diesel | 9 - 87 | 57 - 92 | 29 - 82 |
| Neupane et al.<sup>14</sup> | 20 types | Ambient air particles | 63 - 84 | 94 |  |
| Konda et al.<sup>12</sup> | 15 types | >300 nm | 14 - 99.5 | 99.6 ± 0.1 | 99.9 ± 0.1 |
| | | <300 nm | 9 - 97 | 76±22 | 85±15 |
| Ma et al.<sup>13</sup> | 1 layer of polyester cloth + 4 layers of kitchen paper | Droplets containing Avian Influenza Virus | 95.15 | 97.14 | 99.98 |
|  |  |  | (95% CI 90.97 - 97.39) | (95% CI 94.36 - 98.55) | (95% CI 99.98 - 99.99) |

Almost all studies showed that surgical masks and N95 masks were superior to cloth masks, except one. In the study by Shakya et al.^11^ a certain type of cloth mask performed better than surgical masks and even N95 masks in some cases. N95 also performed poorly in the study by Konda et al.^12^ when the particle size was less than 300 nm.

The study by van der Sande et al.^8^ measured the filtration efficacy of cloth masks on expulsion of droplets by a person and found it to be about 14%.

Filtration efficacy of cloth masks varied depending on the material used. A value as low as 9% was obtained while the highest value was 99.5%. As per the available data from the included studies cloth masks had better filtration efficacy in case of larger particles similar to surgical masks and N95 masks.

In the study by Shakya et al.^11^ one of the cloth masks having an exhaust valve performed exceptionally well in case of particles larger than 1 *μ*m in size. It showed a filtration efficacy of 81% compared to 78% efficacy of the surgical mask.

Konda et al.^12^ assessed the filtration efficacy of 15 natural and synthetic fabrics. The best performers in the <300 nm range were the cotton/chiffon hybrid, cotton quilt, cotton/flannel hybrid and cotton/silk hybrid with above 90% for all of them. In the range of >300 nm however, the best performers with >90% efficacy were the double layered and single layered cotton with 600 threads per inch, cotton/chiffon hybrid, cotton-silk hybrid and cotton quilt.

### Effect of Leakage

Two studies evaluated the effect of leakage around the masks on the filtration capacity. In both studies the filtration efficacy dropped significantly in the presence of leakage.

In 1983 Cooper et al.^7^ observed that fixation by nylon hosiery reduced penetration up to 18% in N95 masks and 40% in cloth masks made of four layers of handkerchief. Konda et al.^12^ also found that in the presence of leakage, the efficacy of all masks decreased by about 50% or more.

### Microscopic Properties

Neupane et al.^14^ examined 20 different types of cloth masks under light microscope and found that the smallest pore size was 81 ± 29 *µ*m, whereas the largest pore size was 461 ± 108 *µ*m. The pore density was found to be 12 to 47 pores / 4.5 mm^2^. The surfaces of the cloth masks were distorted significantly under stretch and the pore size increased, but the surgical masks did not show any such effect. The washing and drying of a cloth mask was also observed to gradually decrease its filtration efficacy (R^2^ = 0.99). After four cycles of washing and drying the filtration efficacy dropped by about 20%.

## Discussion

Available clinical data suggests that cloth masks are inadequate in preventing influenza like illness in healthcare settings. However, no relationship was found between compliance and clinical efficacy. Filtration efficacy of cloth masks was found to be variable between different types of materials used in cloth masks. Filtration efficacy is also reduced significantly by leakage.

The microscopic study^14^ of the cloth masks revealed that the pore size of cloth masks is larger than 50 *µ*m (smallest pore size 81 ± 29 *µ*m). Yang et al.^15^ plotted the size distribution of droplets produced by coughing and found three peaks at 1 *µ*m, 2 *µ*m and 8 *µ*m size bands, which are much smaller than the pores in the cloth masks.

However, the studies on the filtration efficacy of cloth masks report a decent efficiency of cloth masks in filtering out particulate matters for certain types of fabrics. In some cases, the efficacy of cloth masks has been reported to be higher than 90%. In the study by Konda et al.^12^ double layered cotton with 600 threads per inch showed a filtration efficacy of 99.5 ± 0.1% in the >300 nm particle range compared to 99.9 ± 0.1% efficacy of N95 masks in the same range. Although less than the N95 masks in most cases such high efficiency of some types of cloth masks raises the hope of them being useful against droplet infections.

In the filtration efficacy studies the efficiency of the cloth masks in filtering particulate matters remained decent even with the particles smaller than the pore size. This may be due to multiple layers of cloths. The electrostatic forces in the fabric threads may also play a role.

However, cloth masks are not standardized and there is a wide range of cloth masks available with different quality fabrics. Each of these studies used different types of cloth masks. Which may explain the apparent disparity among the study results. The cloth masks are also rarely fit tested which causes significant decrease in efficiency due to leakage, which may be up to 50% or even more.^12^

In the healthcare setting, cloth masks were found to be inadequate^5^ in preventing influenza like illness compared to the surgical masks (relative risk 13.25, 95% confidence interval 1.74 to 100.97). However, the conclusion must be drawn carefully as there was no arm without mask. It is not possible to deduce whether the use of these masks is better than no masks. Moreover the filtration test of the cloth masks in that study showed only 3% efficacy, so the poor performance of the cloth masks may have been due to their quality.

The study on compliance,^6^ however, failed to show any relation between the infection rate and the mask use. This result puts into question whether use of masks has any effect on preventing infection or not. This finding contrasted with a previous Cochrane Review^2^ in 2011. In this review, a meta-analysis of seven case-control studies showed that the odds ratio of mask use vs control was 0.32 (95% confidence interval 0.26 to 0.39) regarding the occurrence of infection by respiratory viruses. But the question of efficacy of masks, as a whole, is beyond the scope this review.

Despite good filtration efficacy, a poor protective effect against the infection by respiratory pathogens may be explained by a number of reasons. First, the leakage around the masks may contribute significantly in reducing the efficacy. In fact, two of the included studies^7,12^ showed that it may reduce the efficacy by as much as 50%. Another possibility is that, despite a decently high efficacy of cloth masks the small amount of particles passing through may be sufficient to cause a clinical infection. However, these issues are also likely to affect the efficacy of medical grade masks.

From the analysis of the available evidence, it is clear that further clinical studies are needed to resolve the apparent disparity. If such a trial is conducted it would be necessary to use different types of cloth masks. There is also a lack of study in the community setting. The only available clinical study was done in a healthcare setting and the data cannot be extrapolated for the use of cloth masks in the community.

During a pandemic the scarcity of resources might prevent the distribution of surgical masks to the community. The use of cloth masks may arise in times like that. As per the available evidences use of cloth masks may be recommended to the general population. However, it must be ensured that the people using the masks understand its limited efficacy against the infection, otherwise they might fall victim to a false sense of security. Use of cloth masks should not lead to a neglect of other infection control measures.

Evidence^10^ shows that cloth masks were also capable of reducing droplet expulsion during coughing compared to the absence of a mask. Hence, maximum safety with cloth masks may be obtained if both the infected and the healthy persons were the masks.

If cloth masks are recommended for community use, it would perhaps be advisable to standardize the masks with usage of the materials proven to have high filtration efficacy. Leakage needs to be minimized as much as possible. The users should also be instructed to use the masks properly and replace them regularly rather than repeatedly using the same mask.

In the healthcare setting, use of cloth masks cannot be recommended. The available evidence indicates that cloth masks are grossly ineffective in preventing infection by respiratory pathogens in the healthcare workers. The standard protocol in this context should include surgical masks and respirators like N95.

Unlike the disposable surgical masks, cloth masks are often washed and dried repeatedly and used for a prolonged period of time. However, the study on the microscopic structure of the cloth masks^14^ showed that repeated washing and drying may reduce the quality of the cloth mask, by almost 20% after four such cycles. In the light of this evidence, it may be advisable to change the cloth masks regularly rather than repeated washing.

Wearing a cloth mask for a prolonged time continuously would also accumulate the respiratory pathogens on the outer surface. This may result in self-contamination from the mask itself. A study on the surgical masks^16^ found that this effect increases with the duration of mask wear. The effect is most likely similar in cloth masks. We need a similar study with the cloth masks to determine their maximum safe period of continuous use.

Our study had several limitations. The number of clinical studies was very small. Also, the results of the included studies were too heterogeneous to allow for a meta-analysis. The paper by Shakya et al.^11^ only had graphical representation of the data. This data had to be extracted by use of a graph digitizer which maybe prone to some degree of inaccuracy. The lack of standardization of the cloth masks meant that every study used different types of masks, which may lead to non-comparable results and make interpretation difficult.

## Conclusion

Although the filtration efficacy of cloth masks is generally lower compared to the surgical masks and N95 masks, they are capable of filtering out some fraction of particles and hence may be better than using no masks at all. In the community setting cloth mask may be recommended during a pandemic caused by respiratory pathogens if medical grade masks are in short supply.

However, the randomized controlled trial showed^5^ that cloth masks are likely to be inadequate in the healthcare setting, so it may be advisable to avoid recommending them to the healthcare workers. However, more randomized controlled trials is needed in both the healthcare and the community settings to generate adequate evidence.

Efficacy of cloth masks varies greatly depending on the materials used, which may be improved by standardization of the manufacturing process of cloth masks. Appropriate instructions for their use, proper fitting to avoid leakage and regular change of cloth masks are essential to maximize their protective efficacy.

## Data Availability

Data regarding study inclusion, data extraction is available as supplementary material.

https://file.agnibho.com/supplementary/systemic-review-cloth-mask-efficacy/

## Author Contributions

AM conceived and prepared the outline of the study. All authors were involved in the study design. AM and AD were involved in the collection of data and served as the first and the second reviewer. RPG acted as the overall supervisor of the study and the third reviewer. The first draft of the manuscript was prepared by AM which was then further edited by RPG and AD.

